# The bidirectional relationship between brain structure and physical activity: a cohort study in the UK Biobank

**DOI:** 10.1101/2023.02.07.23285571

**Authors:** M. Rodriguez-Ayllon, Meike W. Vernooij, Ryan Muetzel, Alexander Neumann, Amy Hofman, Julia Neitzel

## Abstract

Physical activity has been suggested to project again brain atrophy, but also to be a determinant of physical activity in older adults. However, research is needed to confirm this bidirectional relationship. Therefore, this research aimed to study the bidirectional relationship between physical activity and brain structure in older adults from the UK Biobank. A total of 3,027 (62.45 ± 7.27 years old, 51.3% females) had physical activity and MRI data at baseline and follow-up. Physical activity was assessed through a self-reported questionnaire. T1-weighted MRI and diffusion tensor imaging were used to quantify brain volumes and white matter microstructure, respectively. Cross-lagged panel models were performed to estimate bidirectional associations, and linear mixed-effects models (LMM) to investigate the consistency of findings. Overall, our main findings suggest that higher hippocampal volume (Standardized Beta Coefficient (β)=0.075, p_FDR_<0.001), frontal volume (β=0.043, p_FDR_=0.037), and global fractional anisotropy (β=0.042, p_FDR_=0.028) were associated with more do-it-yourself (DIY) activities (e.g., watering the lawn or digging) levels at follow-up. In addition, strenuous sports at baseline were positively associated with hippocampal volume over time (β=0.011, p_FDR_=0.108). Although this association did not survive/was not significant after adjustment for multiple comparisons, was confirmed by the LMM. In contrast, higher levels of walking for pleasure were negatively associated with white matter volume at follow-up (β= −0.026, p_FDR_=0.008). In conclusion, there is a bidirectional association between physical activity and brain structure in healthy middle-aged and older adults. However, further research is needed to understand why physical activity subdomains are associated differently with brain structures and features during aging.

## INTRODUCTION

The proportion of adults aged 60 years and older will increase by over a billion by 2050. Consequently, several adaptations across all sectors of societies (e.g., health and social care) are urgently needed to adapt to and prevent the increasing prevalence of age-rated diseases^1^.

Due to its multisystemic benefits, increasing physical activity levels is considered one of the most promising strategies to prevent diseases, improve quality of life, and reduce public health-care costs in older adults^2–4^. In contrast, the age-related decline in physical and mental capacity leads older adults to avoid physical activity, which results in higher rates of physical inactivity at these ages^4^. Altogether, the limited success in getting and keeping older adults physically active and the urgent need to increase physical activity levels at these ages emphasize the necessity of identifying physical activity determinants in older adults. In this line, previous studies observed that physical activity levels can be predicted by physical health indicators (e.g., physical functioning), other lifestyle behaviors (e.g., no smoking), or psychosocial factors (e.g., self-efficacy)^5^. More recently, it has also been suggested that a healthier brain structure (e.g., gray/white matter volume) predicts a lower decline in physical activity levels in older people^6,7^. In particular, it was observed that larger total brain volume, gray matter volume, and white matter volume were associated with increased sports participation in older people from the Netherlands^6^. Similarly, Arnardottir et al. found that total brain volume was positively associated with physical activity levels in older adults from Iceland^7^. A possible explanation for this might be that a healthier brain structure results in better executive function, which could help older people to discern better between healthy and unhealthy behaviors^8^. For instance, older adults with higher working memory or cognitive flexibility might be more able to plan or prioritize physical activity routines in their daily life.

Even when optimal levels of activity are frequently not achieved by older people, physical activity seems to be associated with healthier cognitive aging in older people^9^, which suggests the association between physical activity and brain structure might be bidirectional. Specifically, several studies have explored the association between physical activity and brain volumes, particularly in the hippocampus^10,11^. However, the literature remains still controversial. For instance, a systematic review, based on observational studies, suggested that physical activity is associated with larger brain volumes (i.e., less brain atrophy)^12^. In contrast, a recently published meta-analysis of experimental studies found no significant effect of exercise interventions on brain volume changes among older adults^13^. With regard to white matter microstructure, a longitudinal study suggested that better maintenance of time spent walking over a decade was associated with slower deterioration in global microstructural features of white matter over time^14^. Contrary, we recently observed that higher levels of physical activity were not associated with better white matter microstructure over time^6^.

Overall, more research is needed to confirm whether poorer brain structure is a risk factor for physical inactivity during aging. The confirmation of those results might help to make policymakers more aware of the need for prioritizing effective interventions in this target group of older people with poor brain health. Additionally, exploring whether a higher volume in brain regions associated with executive function (e.g., frontal lobe) predicts more physical activity over time might shed light on the mechanisms linking brain structure with physical activity in older people. Lastly, identifying which types of daily physical activities protect the brain structure of older people might help to develop effective strategies to delay the neuro decline produced during aging. Therefore, the aim of this study was to study the bidirectional relationship between physical activity and brain structure, while taking account potential differences between various physical activity modalities and brain regions, in a cohort of older adults from the UK

## METHODS

### Study design and participants

This study used data from a large community-based cohort of UK Biobank, which enrolled 502,507 individuals aged 40 and 69 years across the United Kingdom (UK) between 2006 and 2010. The UK Biobank core study comprises >40,000 imaged subjects. Participants were scanned at three centers with identical Siemens Skyra 3T scanners using a standard 32-channel head coil^15^. Imaging was performed 9.60 ± 1.10 years after study baseline. In total, 43,304 participants had physical activity and MRI data at one time point (‘baseline’: 2014+ or ‘follow-up’: 2019+, median duration between visits: 2 years). Of the 43,304 participants, a total of 3,027 had physical activity and MRI data at baseline and follow-up by 07.07.2021 (See **Figure 1**).

**Figure 1.**
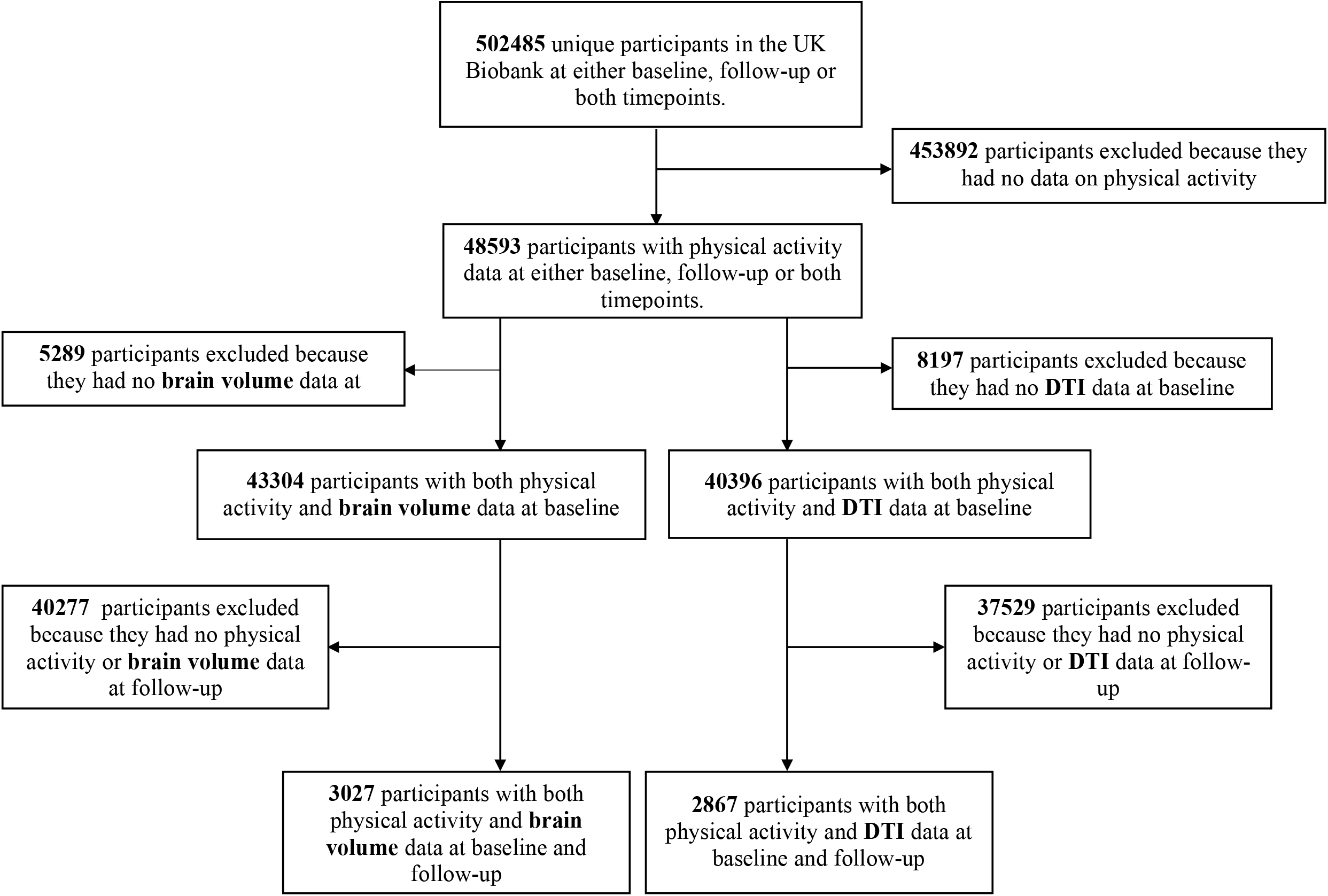
Flow chart. DTI= Diffusion tensor imaging. Participants at follow-up are a subsample, it means they are not part of a drop-out.

### Physical activity

Information on the levels of physical activity was obtained through a self-reported questionnaire administered when the MRI scan. To assess the level of physical activity, respondents indicated the duration (i.e., less than 15 minutes, between 15 and 30 minutes, between 30 minutes and 1 hour, between 1 and 1.5 hours, between 1.5 and 2 hours, between 2 and 3 hours) per the frequency (i.e., once in the last 4 weeks, 2-3 times in the last 4 weeks, once a week, 2-3 times a week, 4-5 times a week, every day) they engage in: walking for pleasure (not as a means of transport), strenuous sports, other exercises (e.g., swimming, cycling, keep fit, bowling), light do-it-yourself (DIY) activities (e.g., pruning, watering the lawn), and heavy DIY activities (e.g., weeding, lawn mowing, carpentry, digging). A total DIY activities score was calculated by adding the hours of light DIY and heavy DIY. In addition, a total physical activity score was calculated by adding the hours of walking for pleasure, strenuous sports, other exercises and DIY activities.

### Magnetic resonance imaging

Overall, T1, T2 FLAIR, and DTI images were used in this analysis. Details on UKB preprocessing and quality control pipelines can be found (<u>HERE</u>).

### Image acquisition and processing

T1-weighted images were obtained using an MPRAGE sequence: TR=2000ms, TE=2.0ms, 208 sagittal slices, flip angle=8°, FOV=256mm, matrix=256×256, slice thickness=1.0mm (voxel size 1×1×1mm). T2-weighted FLAIR imaging was additionally acquired with 3D SPACE in the sagittal plane (resolution = 1.05 × 1 × 1 mm, field of view = 192 × 256 × 256 mm; inversion time = 1800 ms, repetition time = 5000 ms). Diffusion images were obtained using a spin-echo echo-planar sequence with 10 T2-weighted baseline volumes, 50b = 1000 s mm-2 and 50 b=2000 s mm-2 diffusion weighted volumes, with 100 diffusion-encoding directions and 2mm isotropic voxels.

Summary measures of brain structure (i.e., total brain volume, gray matter volume, white matter volume, white matter hyperintensity, hippocampal volume, frontal lobe volume, global fractional anisotropy [FA], global mean diffusivity [MD]) have been generated on behalf of UK Biobank^16^, and are available from UK Biobank upon data access application. Global brain volume measures were normalized for head size. Therefore, we did not adjust our analyses for other volumetric MRI measures such as intracranial volume. For a detailed description of the imaging protocol and pre-processing steps, please see the **Supplementary Material**.

### Covariates

All models were adjusted for age when participants attended for the first time to the MRI assessment center (Field 21003); sex (Field 31); education, categorized as higher (college/university degree or other professional qualification) or lower (Field 6138)^17^; ethnicity (Field 21000), categorized as white/non-white; and body mass index (BMI) at baseline (Field 21001).

Fully adjusted models also included the following covariates assessed at baseline: diet quality (Fields 1309, 1319, 1289, 1299, 1448, 1438, 1468, 1458, 1329, 1339, 1408, 1418, 1428, 2654, 1349, 1359, 1369, 1379,1389, 3680), smoking status (Field 20116), and hypertension (Fields 4079-80), which were defined according to the LS7 score for ideal cardiovascular health^17^. In addition, depression was defined by any of the ICD-10 codes (Field 41270) F32 (depressive episode) or F33 (recurrent depressive disorder). Cardiovascular disease was defined by any of the ICD-10 codes I20-I25 (coronary/ischaemic heart diseases), I46 (cardiac arrest), I48 (atrial fibrillation), I50 (heart failure), I60-I69 (cerebrovascular diseases) as well as algorithmically defined stroke outcomes (ischemic stroke, intracerebral hemorrhage, and subarachnoid hemorrhage; Fields 42006–42013)^18^. Diabetes was defined by ICD-10 codes E10-14 (Diabetes mellitus). Cancer was obtained from the cancer registry records and considering all ICD-10 cancer (‘C’) code entries (Field 40005, 40006, 40008, 40009), except C44 (other malignant neoplasms of skin)^19^.

Lastly, dementia diagnosis was obtained from the algorithmically defined dementia outcomes (all-cause dementia, Alzheimer’s disease, vascular dementia, frontotemporal dementia; Fields 42018-42025) or any of the ICD-10 diagnosis codes F00 (dementia in Alzheimer’s disease), and F01 (vascular dementia). No participants were diagnosed with dementia when baseline and follow-up measures were obtained. Therefore, the dementia variable was not used as a confounder in this study.

### Statistical analysis

The bidirectional associations of total physical activity and the specific domains of physical activity with brain tissue volumes and white matter microstructure were examined using a cross-lagged panel model approach using the Lavaan package in R.^20^

In these path analyses, all associations were adjusted for each other: i.e., analyses are adjusted for the underlying associations of physical activity over time (autoregressive path, β_AR-PA_), brain variables over time (autoregressive path, β_AR-MRI_), the cross-sectional paths (β_CS-Baseline),_ and the prospective mutual associations that represent the bidirectional associations between physical activity and brain variables: the cross-lagged pathways β_CL-1_ and β_CL-2_. These path analyses generate standardized structural regression coefficients (i.e., per standard deviation change) that can be directly compared to assess the direction of the association between physical activity and brain structure^6,21^. To preserve all available data (missing data in baseline covariates ≤ 10%, except for diet quality: 20% and hypertension: 30%), we used the Full Information Maximum Likelihood estimator, as implemented in Lavaan^20^. This is a standard approach to prevent listwise deletion of participants with missing data^20^.

First, we assessed the associations between total physical activity and brain structure adjusted for age, sex, educational level, national origin, and BMI (model 1). In the second model, we additionally adjusted for other behaviors (i.e., diet quality and smoking) and diseases (i.e., hypertension, cancer cardiovascular diseases, diabetes and depression). To adjust for multiple comparisons, we used false discovery rate based on the Benjamini-Hochberg method^22^. We adjusted each pathway for a total of eight tests (i.e., 6 measures of brain volumes and 2 measures of white matter microstructure).

Associations that showed a significant effect (before adjusting for multiple comparisons) in the cross-lagged panel model were re-assessed by using linear mixed-effects models with random intercepts. Follow-up time in years after baseline measurement was used as the time variable, a more detailed description of the linear mixed-effects modeling approach can be found in the **Supplementary Material**. As both modeling types have strengths and limitations, linear mixed-effects models were used to investigate consistency of our findings^23^. Therefore, we considered consistent results across these two methods as most robust. Also, linear mixed-effects models do provide a measure of within-subject change, while cross-lagged models do not without random effects, which require additional time points.

A number of sensitivity analyses were performed. First, we ran the models excluding those people who were less able to perform physical activity (defined as those who were by their doctor restricted in physical activity due to heart condition or feel chest pain felt during physical activity) (Field 6014-6015). Second, a non-linear (quadratic) age term was added to the model in order to ascertain linearity assumptions.

Statistical analyses were performed using R version 4.2.1 (The R Foundation for Statistical Computing, Vienna, Austria).

## RESULTS

The mean age of the study population was 62.45 ± 7.27 years at baseline, and 64.78± 7.22 at follow-up (**Table 1**). 51.3% of the participants were female. At baseline, participants reported a total physical activity of 6.51 ± 5.88 hours per week, of which walking for pleasure was the most prevalent (2.45 ± 3.05 hours per week). At follow-up, participants reported a total physical activity of 6.39 ± 5.95 hours per week. Overall, participants decreased their physical activity levels at follow-up in all domains except for walking for pleasure which increased (see **Table S1**). The subsample of imaged participants had a healthier lifestyle and was less often diagnosed with chronic diseases than the non-imaged participants^24^. Notably, compared with the general population, participants of the UK Biobank were less likely to be obese, smoke, drink alcohol, and report health conditions^25^.

**Table 1.**
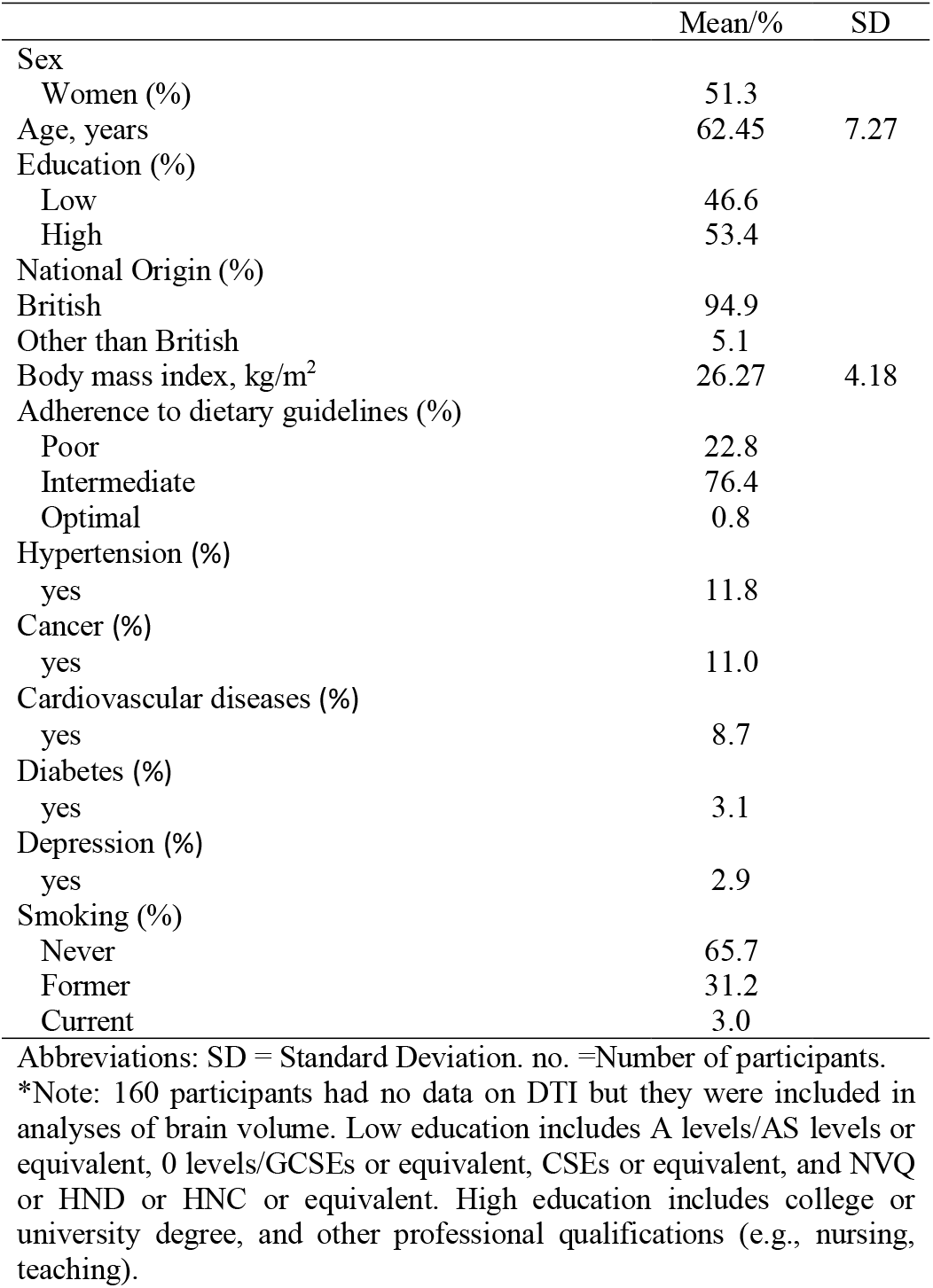
Study sample characteristics at baseline (n=3,027)

|  | Mean/% | SD |
| --- | --- | --- |
| Sex |  |  |
| Women (%) | 51.3 |  |
| Age, years | 62.45 | 7.27 |
| Education (%) |  |  |
| Low | 46.6 |  |
| High | 53.4 |  |
| National Origin (%) |  |  |
| British | 94.9 |  |
| Other than British | 5.1 |  |
| Body mass index, kg/m<sup>2</sup> | 26.27 | 4.18 |
| Adherence to dietary guidelines (%) |  |  |
| Poor | 22.8 |  |
| Intermediate | 76.4 |  |
| Optimal | 0.8 |  |
| Hypertension (%) |  |  |
| yes | 11.8 |  |
| Cancer (%) |  |  |
| yes | 11.0 |  |
| Cardiovascular diseases (%) |  |  |
| yes | 8.7 |  |
| Diabetes (%) |  |  |
| yes | 3.1 |  |
| Depression (%) |  |  |
| yes | 2.9 |  |
| Smoking (%) |  |  |
| Never | 65.7 |  |
| Former | 31.2 |  |
| Current | 3.0 |  |

Abbreviations: SD = Standard Deviation. no. =Number of participants.
\*Note: 160 participants had no data on DTI but they were included in analyses of brain volume. Low education includes A levels/AS levels or equivalent, 0 levels/GCSEs or equivalent, CSEs or equivalent, and NVQ or HND or HNC or equivalent. High education includes college or university degree, and other professional qualifications (e.g., nursing, teaching).

Cross-lagged models exploring the bidirectional association between brain structure variables and total physical activity adjusted for age, sex, educational level, national origin, BMI, other behaviors (i.e., diet quality and smoking), and other diseases (i.e., hypertension, cancer cardiovascular diseases, diabetes, and depression) are presented in **Table 2**. Overall, our results indicated that lower burden of white matter hyperintensity (β=−0.040, p_FDR_=0.016), and greater hippocampal volume (β=0.048, p_FDR_=0.016) at baseline, were associated with higher total physical activity levels at follow-up. Total physical activity at baseline was also associated with larger total gray matter volume at follow-up (β=0.012, p_FDR_=0.033), but this association was not significant after adjustment for multiple comparisons (p=0.264). Results were similar when the model was adjusted for basic confounders (**Table S2**). According to the fit measures (i., comparative fit index and root mean square error of approximation) included in **Table 2** (model 2) and **Table S2** (model 1), the fully adjusted model fit better than the basic one. Therefore, model 2 was considered as the principal analysis, while model 1 results were moved to the supplement.

**Table 2.**
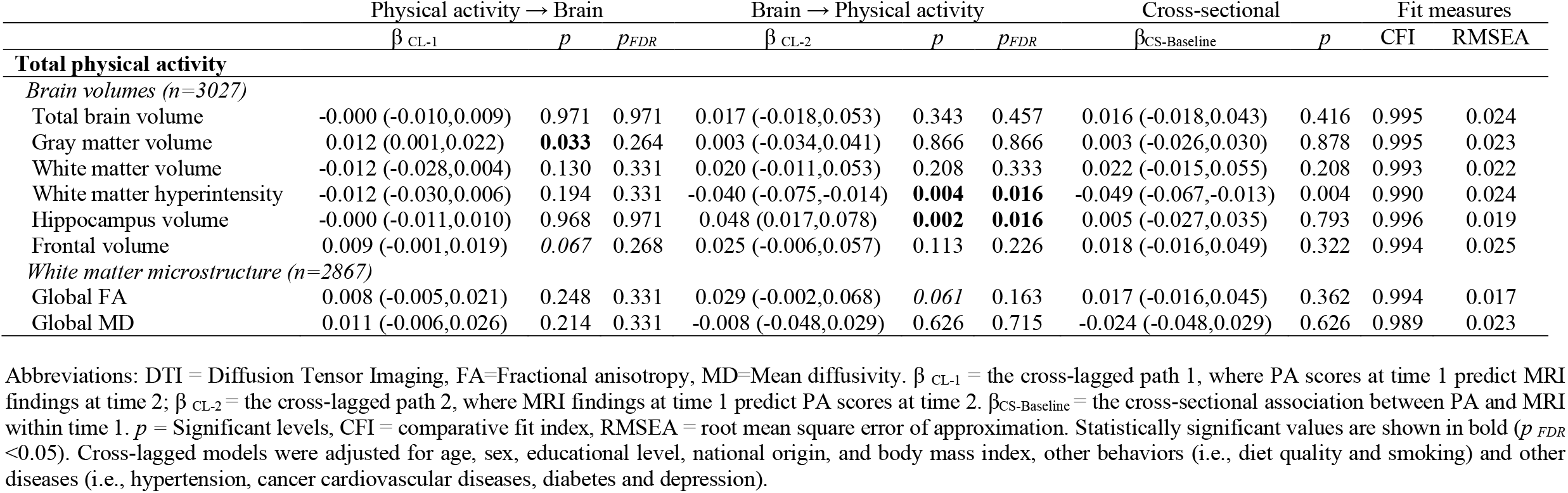
Bidirectional associations between total physical activity and brain structure based on cross-lagged panel models.

Associations of specific physical activity domains and brain structure indicators adjusted for age, sex, educational level, national origin, BMI, other behaviors (i.e., diet quality and smoking), and other diseases (i.e., hypertension, cancer cardiovascular diseases, diabetes, and depression) are presented in **Table 3**. Contrary to our hypothesis, higher levels of walking for pleasure at baseline were negatively associated with white matter volume at follow-up (β=−0.026, p_FDR_=0.008). Interestingly, a positive association of strenuous sports with hippocampal volume (β=0.011, p=0.027) and frontal volume (β=0.011, p=0.013) was observed, although these associations disappeared after adjusting for multiple comparisons (p_FDR_>0.1). Results were similar when the model was adjusted for basic confounders (see **Table S3**).

**Table 3.** Bidirectional associations between physical activity domains and brain structure based on cross-lagged panel models (model 2).

|  | Physical activity → Brain |  |  | Brain → Physical activity |  |  | Cross-sectional |  | Fit measures |  |
| --- | --- | --- | --- | --- | --- | --- | --- | --- | --- | --- |
| | $\beta_{CL-1}$ | $p$ | $p_{FDR}$ | $\beta_{CL-2}$ | $p$ | $p_{FDR}$ | $\beta_{CS-Baseline}$ | $p$ | CFI | RMSEA |
| **Walking for pleasure** |  |  |  |  |  |  |  |  |  |  |
| *Brain volumes (n=3027)* |  |  |  |  |  |  |  |  |  |  |
| Total brain volume | -0.011 (-0.020,-0.001) | **0.026** | 0.104 | 0.041 (0.006,0.076) | **0.023** | 0.184 | -0.003 (-0.032,0.028) | 0.883 | 0.996 | 0.022 |
| Gray matter volume | 0.007 (-0.004,0.017) | 0.244 | 0.437 | 0.036 (-0.001,0.074) | 0.056 | 0.224 | -0.018 (-0.041,0.014) | 0.339 | 0.996 | 0.021 |
| White matter volume | -0.026 (-0.041,-0.010) | **0.001** | **0.008** | 0.026 (-0.004,0.056) | 0.090 | 0.236 | 0.013 (-0.022,0.046) | 0.495 | 0.995 | 0.020 |
| White matter hyperintensity | -0.013 (-0.030,0.005) | 0.146 | 0.389 | -0.020 (-0.049,0.006) | 0.118 | 0.236 | 0.036 (-0.002,0.043) | 0.070 | 0.913 | 0.057 |
| Hippocampus volume | 0.004 (-0.007,0.015) | 0.502 | 0.564 | 0.015 (-0.015,0.043) | 0.336 | 0.384 | 0.078 (0.011,0.035) | <0.001 | 0.939 | 0.063 |
| Frontal volume | 0.005 (-0.005,0.015) | 0.335 | 0.447 | -0.003 (-0.032,0.026) | 0.839 | 0.839 | 0.020 (-0.005,0.017) | 0.307 | 0.949 | 0.057 |
| *White matter microstructure (n=2867)* |  |  |  |  |  |  |  |  |  |  |
| Global FA | 0.004 (-0.010,0.018) | 0.564 | 0.564 | 0.023 (-0.009,0.060) | 0.152 | 0.243 | 0.061 (0.021,0.082) | 0.001 | 0.996 | 0.014 |
| Global MD | 0.010 (-0.008,0.028) | 0.273 | 0.437 | -0.018 (-0.058,0.016) | 0.270 | 0.360 | -0.048 (-0.067,-0.009) | 0.011 | 0.992 | 0.020 |
| **Other exercises** |  |  |  |  |  |  |  |  |  |  |
| *Brain volumes (n=3027)* |  |  |  |  |  |  |  |  |  |  |
| Total brain volume | -0.003 (-0.012,0.007) | 0.622 | 0.784 | -0.004 (-0.037,0.030) | 0.825 | 0.861 | -0.003 (-0.032,0.026) | 0.844 | 0.997 | 0.019 |
| Gray matter volume | 0.001 (-0.009,0.011) | 0.838 | 0.838 | -0.019 (-0.056,0.017) | 0.292 | 0.467 | 0.011 (-0.017, 0.033) | 0.520 | 0.997 | 0.019 |
| White matter volume | -0.006 (-0.021,0.010) | 0.484 | 0.784 | 0.010 (-0.019,0.040) | 0.490 | 0.653 | -0.016 (-0.049,0.019) | 0.380 | 0.996 | 0.017 |
| White matter hyperintensity | -0.012 (-0.028,0.006) | 0.190 | 0.507 | -0.019 (-0.050,0.007) | 0.134 | 0.357 | -0.037 (-0.056,-0.006) | 0.015 | 0.993 | 0.019 |
| Hippocampus volume | -0.003 (-0.015,0.010) | 0.686 | 0.784 | -0.016 (-0.046,0.013) | 0.285 | 0.467 | -0.014 (-0.043,0.018) | 0.433 | 0.998 | 0.014 |
| Frontal volume | 0.008 (-0.004,0.018) | 0.188 | 0.507 | 0.003 (-0.027,0.032) | 0.861 | 0.861 | 0.010 (-0.025,0.043) | 0.609 | 0.995 | 0.021 |
| *White matter microstructure (n=2867)* |  |  |  |  |  |  |  |  |  |  |
| Global FA | 0.013 (-0.001,0.026) | 0.073 | 0.507 | -0.034 (-0.074,-0.006) | **0.021** | 0.168 | -0.001 (-0.032,0.031) | 0.964 | 0.998 | 0.011 |
| Global MD | 0.005 (-0.013,0.021) | 0.616 | 0.784 | 0.025 (-0.003,0.065) | 0.076 | 0.304 | -0.008 (-0.036, 0.023) | 0.660 | 0.994 | 0.018 |
| **Strenuous sports** |  |  |  |  |  |  |  |  |  |  |
| *Brain volumes (n=3027)* |  |  |  |  |  |  |  |  |  |  |
| Total brain volume | 0.005 (-0.003,0.012) | 0.255 | 0.510 | 0.015 (-0.021,0.052) | 0.409 | 0.628 | 0.013 (-0.018,0.039) | 0.459 | 0.995 | 0.021 |
| Gray matter volume | 0.000 (-0.008,0.008) | 0.939 | 0.939 | 0.010 (-0.026,0.048) | 0.550 | 0.628 | -0.006 (-0.030,0.021) | 0.726 | 0.996 | 0.022 |
| White matter volume | 0.009 (-0.002,0.020) | 0.109 | 0.291 | 0.011 (-0.021,0.044) | 0.481 | 0.628 | 0.026 (-0.008,0.058) | 0.144 | 0.994 | 0.020 |
| White matter hyperintensity | 0.003 (-0.011,0.017) | 0.693 | 0.924 | -0.013 (-0.044,0.015) | 0.336 | 0.628 | -0.009 (-0.025,0.009) | 0.372 | 0.990 | 0.023 |
| Hippocampus volume | 0.011 (0.001,0.019) | **0.027** | 0.108 | 0.036 (0.004,0.070) | **0.027** | 0.216 | 0.011 (-0.019,0.039) | 0.497 | 0.997 | 0.017 |
| Frontal volume | 0.011 (0.002,0.018) | **0.013** | 0.104 | 0.023 (-0.009,0.058) | 0.156 | 0.624 | 0.005 (-0.029,0.038) | 0.793 | 0.994 | 0.023 |
| *White matter microstructure (n=2867)* |  |  |  |  |  |  |  |  |  |  |
| Global FA | 0.001 (-0.012,0.014) | 0.848 | 0.939 | 0.004 (-0.035,0.045) | 0.801 | 0.801 | -0.007 (-0.035,0.024) | 0.709 | 0.995 | 0.015 |
| Global MD | 0.005 (-0.011,0.021) | 0.551 | 0.882 | 0.018 (-0.018,0.064) | 0.271 | 0.628 | 0.005 (-0.023,0.031) | 0.781 | 0.990 | 0.021 |
| **DIY activities** |  |  |  |  |  |  |  |  |  |  |
| *Brain volumes (n=3027)* |  |  |  |  |  |  |  |  |  |  |
| Total brain volume | 0.008 (-0.003,0.018) | 0.147 | 0.588 | -0.009 (-0.048,0.029) | 0.634 | 0.689 | 0.023 (-0.014,0.051) | 0.259 | 0.997 | 0.017 |
| Gray matter volume | 0.012 (-0.000,0.023) | 0.052 | 0.416 | -0.024 (-0.063,0.014) | 0.215 | 0.344 | 0.014 (-0.021,0.040) | 0.524 | 0.997 | 0.016 |
| White matter volume | 0.002 (-0.015,0.019) | 0.829 | 0.829 | 0.007 (-0.028,0.043) | 0.689 | 0.689 | 0.023 (-0.014,0.058) | 0.228 | 0.997 | 0.015 |
| White matter hyperintensity | -0.003 (-0.022,0.017) | 0.781 | 0.829 | -0.030 (-0.066,-0.000) | **0.046** | 0.092 | 0.037 (-0.001,0.044) | 0.065 | 0.903 | 0.056 |
| Hippocampus volume | -0.005 (-0.015,0.005) | 0.330 | 0.829 | 0.075 (0.038,0.107) | **<0.001** | **<0.001** | 0.017 (-0.016,0.046) | 0.354 | 0.999 | 0.007 |
| Frontal volume | 0.002 (-0.008,0.013) | 0.638 | 0.829 | 0.043 (0.009,0.077) | **0.014** | **0.037** | 0.022 (-0.012,0.053) | 0.218 | 0.996 | 0.019 |
| *White matter microstructure (n=2867)* |  |  |  |  |  |  |  |  |  |  |
| Global FA | 0.001 (-0.010,0.013) | 0.821 | 0.829 | 0.042 (0.013,0.081) | **0.007** | **0.028** | -0.019 (-0.048,0.016) | 0.321 | 0.999 | 0.007 |
| Global MD | 0.004 (-0.011,0.018) | 0.619 | 0.829 | -0.013 (-0.056,0.025) | 0.454 | 0.605 | 0.005 (-0.030,0.038) | 0.816 | 0.993 | 0.017 |

Abbreviations: DTI = Diffusion Tensor Imaging, DIY= do-it-yourself, FA=Fractional anisotropy, MD=Mean diffusivity. $\beta_{CL-1}$ = the cross-lagged path 1, where PA scores at time 1 predict MRI findings at time 2; $\beta_{CL-2}$ = the cross-lagged path 2, where MRI findings at time 1 predict PA scores at time 2. $\beta_{CS-Baseline}$ = the cross-sectional association between PA and MRI within time 1. $p_{FDR}$ = Significant levels, CFI = comparative fit index, RMSEA = root mean square error of approximation. Statistically significant values are shown in bold ( $p_{FDR} < 0.05$ ). Cross-lagged models were adjusted for age, sex, national origin, educational level, body mass index, other behaviors (i.e., diet quality and smoking) and other diseases (i.e., hypertension, cancer cardiovascular diseases, diabetes and depression).

Some brain structure variables at baseline were also associated with physical activity domains at follow-up. Particularly, our results indicated that larger hippocampal volume (β=0.075, p_FDR_<0.001), frontal volume (β=0.043, p_FDR_=0.037), and global FA (β=0.042, p_FDR_=0.028) were positively associated with higher DIY activities levels at follow-up (**Table 3**). Results were similar when using Model 1 (**Table S3**). Autoregressive coefficients are shown for total physical activity and physical activity domains in **Table S4** and **Table S5**, respectively.

Linear mixed-effects models are presented in **Table S6**. Again, contrary to our hypothesis, linear mixed-effects models confirmed that higher levels of walking for pleasure at baseline predicted a higher decrease in white matter volume over time (β=−0.017, Std.error=0.004). In contrast, higher level of strenuous sports predicted smaller decreases of hippocampal volume over time (β=0.010, Std.error=0.003).

Linear mixed-effects model also confirmed that brain structure predicted physical activity over time (**Table S6**). First, lower white matter hyperintensity predicted lower decreases in DIY activities over time (β=−0.021, Std.error=0.010). Second, higher total brain volume predicted smaller decreases in walking for pleasure over time (β=0.017, Std.error=0.007). Third, higher hippocampal volume predicted smaller decreases in total physical activity and DIY activities (β=0.026, Std.error=0.009). Lastly, higher global FA predicted smaller decreases in DIY (β=0.025, Std.error=0.010), but higher decreases in other exercises (β=−0.019, Std.error=0.008).

Results were similar when less able people to perform physical activity (defined as those whose doctors restricted physical activity due to heart condition or feel chest pain felt during physical activity) were excluded (**Table S7**), or when a non-linear (quadratic) age term was added to the model (**Table S8**).

## DISCUSSION

### Main findings

This study aimed to explore the bidirectional relationship between physical activity and brain structure in a cohort of older adults from the UK. Overall, it seems there is a bidirectional association between physical activity and brain structure in older adults. However, our findings suggest the association might be more consistent when brain structure predicts physical activity levels rather than the reverse association (physical activity predicts brain structure). Notably, the association between brain structure and physical activity levels seems to be driven by DIY activities, which suggests people with a healthier brain structure are more able to deal with their active daily routines. In line with previous literature, we also observed that higher levels of strenuous sports predicted a lower decrease in hippocampal volume over time. Lastly, there was a paradoxical, consistent association between walking for pleasure and white matter volume decrease that requires further investigation.

### The association of brain structure with physical activity over time

Brain structure predicts total physical activity levels over time, and this association seems to be driven by DIY activities. Accordingly, we previously observed that healthier brain structure, in terms of larger brain volume(s) and better white matter integrity, was associated with more self-reported physical activity (i.e., sports and walking) in middle-aged and older adults from the Rotterdam Study^6^. Interestingly, participants from the UK Biobank were on average healthier than those from the Rotterdam Study. For instance, in the present study, only 12% of the participants had hypertension vs. 65% in the Rotterdam Study. High blood pressure is one of the stronger risk factors associated with global and regional brain atrophy in the elderly population^26,27^, which suggests brain structure appears to predict physical activity levels in older people from the general population at different levels of health condition. In addition, it is important to note that both studies included self-reported physical activity, which could under- or overestimations physical activity levels. However, our results are also consistent with those from Arnardottir et al.^7^, who observed that higher grey and white matter volumes were associated with more objectively measured total physical activity in 352 older adults, even when adjusted for self-reported physical activity. Lastly, intervention studies have shown that those participants with a healthier brain structure in terms of brain volume had better adherence to a structured physical activity intervention^28,29^. Altogether previous and current results seem to indicate that brain structure predicts levels of daily movement over time in older adults independently of the physical activity tool (i.e., self-reported vs. objectively measured physical activity) or the structure of the physical activity practice (i.e., DIY activities or structured physical activity interventions). Therefore, future policies designed to keep older adults physically active might consider brain structure (an early indicator of cognitive decline) as a potential physical activity determinant.

### The association of physical activity levels with brain structure over time

This study confirms that higher levels of sports participation are associated with higher volume in the hippocampus of middle aged and older adults. The hippocampus, a subcortical brain structure implicated in memory, spatial navigation, and other aspects of cognitive functioning, is structurally sensitive to exposure and engagement with novel experiences and environments ^30^. It is therefore not surprising that the hippocampus is the most widely studied region consistently associated with physical activity across lifespan^9,11,31–34^. However, the majority of the prior research on physical activity and the hippocampus relies mostly on animal models^35,36^ or clinical samples^37^, had a cross-sectional design^31^, or explored the effect of a structured exercise intervention on the hippocampal volume in late adulthood^34,38^. Interestingly, the only study that has previously explored the bidirectional relationship between unstructured physical activity (free time or self-selected free physical activity) and hippocampal volume in middle-aged and older adults did not observe any association^6^. There are, however, several possible explanations. First, it could be possible physical activity may be less influential in individuals whose brain decline reached a certain threshold^39^. In this sense, differences in the health status of participants from the UK Biobank and the Rotterdam Study might explain why we observed an association only in the UK Biobank sample. Second, other potential reasons, such as differences in the physical activity self-reports or the sample size, might also be considered. Overall, further studies are needed to disambiguate populations most affected by physical activity and the key stages to appreciate brain structure changes (e.g., cognitively normal vs. cognitively impaired) in middle-aged and older adults.

Surprisingly, higher levels of walking for pleasure were associated with lower white matter volume in middle-aged and older adults. Those results were not observed with white matter microstructure (i.e., global FA and global MD) but were consistent in our cross-lagged and linear mixed models. In our previous research from the Rotterdam Study, we also identified an inverse association between walking and white matter volume. However, white matter volume predicted walking rather than the other way around (as observed in the present study). This rather contradictory result is difficult to explain, but there are several possible explanations. For instance, it could be possible that the cognitive demand of the walks, freely chosen by older people, is clouding or confounding the association between walking time and brain structure at these ages. In particular, several factors (e.g., green vs. urban spaces, different vs. same route choosing, walking in group vs. alone) ^40–42^, associated with a higher cognitively enriched walking, could be interacting in the relationship between walking for pleasure and brain structure and might be, then, further explored. Another possible explanation for this could be that not only time dedicated to walking matters but also the way older people walk. In this line, a recent systematic review concluded strong evidence indicated that slower gait speed predicts higher cognitive decline in older people^43^. Overall, further studies are needed to investigate the reasons walking was negatively associated with white matter volume in older adults.

## Strengths and limitations

Several potential limitations should be noted. First, a highly selected sample of healthy participants from the UK was included in this study, which may affect generalizability to the general population. In addition, given that the majority of participants were of Caucasian ethnicity and/or European ancestry, generalizability to other ethnicities needs to be addressed in future studies. Second, physical activity was assessed using a self-report questionnaire that has not been validated and which in turn could overestimate or underestimate the levels of physical activity. This issue could be particularly problematic for physical activity subdomains that do not have a clear start/end (e.g., DIY activities or walking for pleasure), or subdomains such as “other exercises” that include a broad set of heterogeneous activities (i.e., other exercise that could include from bowling to swimming). Third, the observational design limits inferences about causality, and residual confounding cannot be ruled out. However, the confounders that we have measured did not substantially change the estimates of interest, suggesting that the potential for residual confounding might be relatively low. Lastly, we restricted our study to mainly global brain structure metrics. Therefore, we did not capture associations in more specific brain regions. Considering aging is not uniform across the brain, and physical activity could differently affect brain regions, our global approach is a limitation of our study. Nonetheless, the current study took a global approach, in addition to regions well-known to be plastic (i.e., hippocampus), to reduce the number of tests run. The strengths of the current study are the large sample size and the prospectively-collected data across 2 time points. The population-based sampling offers increased generalizability of the findings to a real life setting and physical activity over wide intensity levels compared to previous intervention studies.

### Practical implications

In 2030, 499·2 million new cases of preventable major non-communicable diseases and mental health conditions would occur globally if the prevalence of physical inactivity does not change^44^. In this context, older adults have fewer opportunities to access safe, affordable, and appropriate physical activity programs, missing out on the physical, mental, and social health benefits of being active^45^. Overall, we confirmed the novel findings that suggested poorer brain structure as a potential risk factor for physical inactivity in middle-aged and older people. This new research might help policymakers to prioritize the development of policy actions to promote and enable more middle-aged and older adults with potentially accelerated brain aging to be active. In addition, we have identified higher levels of sports participation and no other types of unstructured physical activities associated with higher volume in the hippocampus over time in healthy middle-aged and older adults. Whole brain and hippocampal atrophy can occur several decades before the onset of cognitive impairments ^46^. Therefore, our study suggests that sport-based physical activity interventions might help to protect the brain health of older adults in an early stage of aging before any apparent cognitive decline.

## Conclusions

There is a bidirectional association between physical activity and brain structure in healthy middle aged and older adults. However, the association seems to be stronger when brain structure predicts physical activity levels rather than the reverse association (physical activity predicts brain structure). Notably, the association between brain structure and physical activity levels seems to be driven by DIY activities. In line with previous literature, we also observed that higher levels of strenuous sports predicted a lower decrease in hippocampal volume over time. Lastly, there was a paradoxical association between walking for pleasure and white matter volume that requires further investigation.

## Data Availability

All data analysed herein (including IDPs) were provided by UK Biobank under project reference 68400, subject to a data transfer agreement. Researchers can apply to use the UK Biobank data resource for health related research in the public interest. A guide to access is available from the UK Biobank website (http://www.ukbiobank.ac.uk/register-apply/).

## Funding

MRA received funding from the Ramón Areces Foundation.

## Conflicts of interest

None.

## Notes

### Competing Interest Statement

The authors have declared no competing interest.

### Funding Statement

MRA received funding from the Ramon Areces Foundation.

### Author Declarations

The study used ONLY openly available human data that were originally located at: the UK Biobank data resource for health related research in the public interest. A guide to access is available from the UK Biobank website (http://www.ukbiobank.ac.uk/register-apply/).

