## Appendix 1 for "The bidirectional relationship between brain structure and physical activity: a cohort study in the UK Biobank"

**Supplementary Material 1. Imaging protocol and pre-processing steps.**

*Total brain volumes*

The T1-weighted images were first defaced to anonymize the images. Second, voxels containing non-brain tissue were removed, using FSL's Brain Extraction Tool[1]; linear registration[2,3], and the MNI152 "nonlinear 6th generation" standard space T1 template (http://www.bic.mni.mcgill.ca/ServicesAtlases/ICBM152NLin6). At this point, gradient distortion correction was also applied. Third, nonlinear registration was executed with FNIRT[4] to calculate the T1-to-MNI152 warp transform, with a custom brain mask as a reference image. Next, the standard space brain mask was transformed into each individual's native T1 space in order to brain extract for each individual using the inverse of the warp transform. Then, FAST[5] was subsequently applied to segment the images based on tissue types (i.e., total brain volume, grey matter volume, white matter volume). Lastly, the pre-processed T1 images were separately submitted to a SIENAX (Structural Image Evaluation, using Normalisation, of Atrophy[1] to derive an estimate of head size (used as a co-variate in the present analyses).

*White matter hyperintensity*

The T2 FLAIR structural images were registered to the T1-weighted images using FLIRT[6]. Then, the transforms generated were used to register the FLAIR images to MNI space. Finally, an estimate of total white matter hyperintensity was generated by feeding the T2-weighted FLAIR and T1-weighted images to the BIANCA tool developed by Griffanti et al.[7]

*Frontal volume*

Grey matter volume of the frontal lobe was estimated from FAST using a combination of the Harvard-Oxford cortical atlases (https://fsl.fmrib.ox.ac.uk/fsl/fslwiki/Atlases) and Diedrichsen cerebellar atlas (http://www.diedrichsenlab.org/imaging/propatlas.htm) to facilitate parcellation.

*Hippocampal volume*

The T1 images were processed using FreeSurfer. The hippocampal region was extracted using FreeSurfer's aseg tool[8]. Next, the FreeSurfer output was quality control checked. In particular, we used the Qoala-T approach[9]. Of note, FreeSurfer output failing this quality control was not included in the analyses.

*White matter microstructure*

First, EPI distortions and eddy currents were addressed with FSl's topup[10] and eddy, respectively[11] for the raw diffusion-weighted images. Second, we applied gradient distortion correction (developed by HCP and FSL) to remove artefacts introduced by head motion. Then, DTI fit[12] was applied to derive FA and MD images for each participant. Finally, the pre-processed diffusion-weighted images were subsequently submitted to a tractography-based analysis. Specifically, BEDPOSTX (Bayesian Estimation of Diffusion Parameters Obtained using Sampling Techniques, http://fsl.fmrib.ox.ac.uk/fsl/fslwiki/FDT/UserGuide) was used. The output of BEDPOSTX were then fed to PROBTRACKX[13–15], a tool that conducts probabilistic tractography, which is able to map 27 major tracts using start/stop ROI masks defined by AutoPX[16].

Global measures of FA and MD were calculated by averaging the diffusion metrics over all white matter tracts of both hemispheres per individual.

**Supplementary Material 2. Description of linear mixed-effects models.**

Linear mixed-effects models were used to investigate consistency of findings, and to obtain a measure of within-subject change. We used linear mixed-effects models with random intercepts, and accounted for different durations between the two timepoints by including follow-up time as the time variable.

Linear mixed-effects models do account for missing values on timepoints for the outcome values. Missing values on baseline covariates were imputed using the 'mice' package in R. We created 30 imputed datasets. Linear-mixed effect models were constructed in each dataset separately and then, effect estimates and p-values were pooled.

Models were fit using the 'lme4' package in R. These models were used to investigate associations between brain metrics at baseline and physical activity over time, i.e., the associations that were significant in cross-lagged panel models. We constructed the models according to the following strategy:

Physical activity $\text{variable}_{ij}$ ~ *Follow-up* $\textit{time}_{ij}$* *Brain structure* $\textit{variable}_{i}$ + *Baseline* $\textit{covariates}_{i}$ + (1 | subject), data=data)

i refers to subject, j refers to timepoint, (1 | subject) refers to the random intercept.

**Table S1.** Physical activity characteristics at baseline and follow-up (n=3,027)

| | Mean/% | SD |
|---|---|---|
| Total physical activity (h/w) | | |
| Baseline | 6.51 | 5.88 |
| Follow-up | 6.39 | 5.95 |
| Walking for pleasure (h/w) | | |
| Baseline | 2.45 | 3.05 |
| Follow-up | 2.58 | 3.25 |
| Strenuous sports (h/w) | | |
| Baseline | 0.36 | 1.23 |
| Follow-up | 0.35 | 1.20 |
| Other exercises (h/w) | | |
| Baseline | 1.46 | 2.17 |
| Follow-up | 1.42 | 2.21 |
| DIY activities (h/w) | | |
| Baseline | 2.24 | 3.96 |
| Follow-up | 2.03 | 3.75 |

Abbreviations: DIY= do-it-yourself ; SD = Standard Deviation; h/w= hours per week. Total physical activity was calculated by adding the hours per week of walking for pleasure, strenuous sports, other exercises and DIY activities.

**Table S2.** Bidirectional associations between total physical activity and brain structure based on cross-lagged panel models.

| | Physical activity → Brain | | | Brain → Physical activity | | | Cross-sectional | | Fit measures | |
|---|---|---|---|---|---|---|---|---|---|---|
| | $\beta_{CL-1}$ | $p$ | $p_{FDR}$ | $\beta_{CL-2}$ | $p$ | $p_{FDR}$ | $\beta_{CS-Baseline}$ | $p$ | CFI | RMSEA |
| **Total physical activity** | | | | | | | | | | |
| *Brain volumes (n=3027)* | | | | | | | | | | |
| Total brain volume | -0.000 (-0.010,0.009) | 0.972 | 0.972 | 0.017 (-0.018,0.053) | 0.344 | 0.459 | 0.018 (-0.016,0.046) | 0.873 | 0.996 | 0.034 |
| Gray matter volume | 0.012 (0.001,0.022) | **0.033** | 0.264 | 0.003 (-0.034,0.041) | 0.869 | 0.869 | 0.007 (-0.023,0.033) | 0.736 | 0.995 | 0.034 |
| White matter volume | -0.012 (-0.028,0.004) | 0.130 | 0.331 | 0.020 (-0.011,0.053) | 0.207 | 0.331 | 0.022 (-0.014,0.056) | 0.248 | 0.996 | 0.028 |
| White matter hyperintensity | -0.012 (-0.030,0.006) | 0.195 | 0.331 | -0.040 (-0.075,-0.014) | **0.004** | **0.032** | -0.056 (-0.073,-0.019) | 0.001 | 0.994 | 0.027 |
| Hippocampus volume | 0.008 (-0.024,0.038) | 0.658 | 0.752 | 0.031 (-0.001,0.016) | *0.084* | 0.224 | 0.008 (-0.024,0.038) | 0.658 | 0.997 | 0.025 |
| Frontal volume | 0.009 (-0.001,0.019) | *0.068* | 0.272 | 0.025 (-0.006,0.057) | 0.113 | 0.226 | 0.022 (-0.013,0.053) | 0.234 | 0.992 | 0.039 |
| *White matter microstructure (n=2867)* | | | | | | | | | | |
| Global FA | 0.008 (-0.005,0.021) | 0.248 | 0.331 | 0.029 (-0.002,0.068) | *0.061* | 0.224 | 0.018 (-0.016,0.045) | 0.349 | 0.955 | 0.024 |
| Global MD | 0.011 (-0.006,0.026) | 0.214 | 0.331 | -0.008 (-0.048,0.029) | 0.626 | 0.715 | -0.025 (-0.050,0.011) | 0.210 | 0.991 | 0.032 |

Abbreviations: DTI = Diffusion Tensor Imaging, FA=Fractional anisotropy, MD=Mean diffusivity. $\beta_{CL-1}$ = the cross-lagged path 1, where PA scores at time 1 predict MRI findings at time 2; $\beta_{CL-2}$ = the cross-lagged path 2, where MRI findings at time 1 predict PA scores at time 2. $\beta_{CS-Baseline}$ = the cross-sectional association between PA and MRI within time 1. $p$ = Significant levels, CFI = comparative fit index, RMSEA = root mean square error of approximation. Statistically significant values are shown in bold ($p_{FDR}$ <0.05). Cross-lagged models were adjusted for age, sex, educational level, national origin, and body mass index.

**Table S3.** Bidirectional associations between physical activity domains and brain structure based on cross-lagged panel models.

| | Physical activity → Brain | | | Brain → Physical activity | | | Cross-sectional | | Fit measures | |
|---|---|---|---|---|---|---|---|---|---|---|
| | $\beta_{CL-1}$ | *p* | $p_{FDR}$ | $\beta_{CL-2}$ | *p* | $p_{FDR}$ | $\beta_{CS-Baseline}$ | *p* | CFI | RMSEA |
| **Walking for pleasure** | | | | | | | | | | |
| *Brain volumes (n=3027)* | | | | | | | | | | |
| Total brain volume | -0.011 (-0.019,-0.001) | **0.026** | 0.104 | 0.041 (0.006,0.076) | **0.024** | 0.192 | -0.002 (-0.032,0.029) | 0.932 | 0.996 | 0.030 |
| Gray matter volume | 0.007 (-0.004,0.017) | 0.240 | 0.437 | 0.036 (-0.001,0.074) | *0.056* | 0.224 | -0.016 (-0.040,0.015) | 0.388 | 0.996 | 0.029 |
| White matter volume | -0.026 (-0.041,-0.010) | **0.001** | **0.008** | 0.026 (-0.004,0.056) | 0.090 | 0.238 | 0.013 (-0.023,0.047) | 0.492 | 0.997 | 0.023 |
| White matter hyperintensity | -0.013 (-0.030,0.004) | 0.145 | 0.387 | -0.020 (-0.049,0.006) | 0.119 | 0.238 | 0.036 (-0.002,0.043) | 0.070 | 0.917 | 0.083 |
| Hippocampus volume | 0.004 (-0.007,0.015) | 0.502 | 0.564 | 0.015 (-0.015,0.043) | 0.336 | 0.384 | 0.078 (0.011,0.035) | <0.001 | 0.940 | 0.094 |
| Frontal volume | 0.005 (-0.005,0.015) | 0.332 | 0.443 | -0.003 (-0.032,0.026) | 0.838 | 0.838 | 0.020 (-0.005,0.017) | 0.304 | 0.949 | 0.087 |
| *White matter microstructure (n=2867)* | | | | | | | | | | |
| Global FA | 0.004 (-0.010,0.018) | 0.564 | 0.564 | 0.023 (-0.009,0.060) | 0.152 | 0.243 | 0.062 (0.022,0.083) | 0.001 | 0.974 | 0.017 |
| Global MD | 0.010 (-0.008,0.028) | 0.273 | 0.437 | -0.018 (-0.058,0.016) | 0.270 | 0.360 | -0.049 (-0.067,-0.009) | 0.011 | 0.969 | 0.026 |
| **Other exercises** | | | | | | | | | | |
| *Brain volumes (n=3027)* | | | | | | | | | | |
| Total brain volume | -0.003 (-0.012,0.007) | 0.626 | 0.788 | -0.004 (-0.037,0.030) | 0.822 | 0.858 | -0.002 (-0.031,0.027) | 0.900 | 0.995 | 0.034 |
| Gray matter volume | 0.001 (-0.009,0.011) | 0.831 | 0.831 | -0.019 (-0.056,0.017) | 0.292 | 0.467 | 0.012 (-0.016, 0.034) | 0.492 | 0.997 | 0.028 |
| White matter volume | -0.006 (-0.021,0.010) | 0.482 | 0.789 | 0.010 (-0.019,0.040) | 0.488 | 0.651 | -0.016 (-0.049,0.019) | 0.388 | 0.998 | 0.020 |
| White matter hyperintensity | -0.012 (-0.028,0.005) | 0.186 | 0.504 | -0.019 (-0.050,0.007) | 0.133 | 0.355 | -0.040 (-0.059,-0.008) | 0.009 | 0.997 | 0.020 |
| Hippocampus volume | -0.003 (-0.014,0.010) | 0.690 | 0.788 | -0.016 (-0.046,0.013) | 0.284 | 0.467 | -0.010 (-0.040,0.022) | 0.565 | 0.999 | 0.018 |
| Frontal volume | 0.008 (-0.004,0.018) | 0.189 | 0.504 | 0.003 (-0.027,0.032) | 0.858 | 0.858 | 0.013 (-0.022,0.046) | 0.491 | 0.995 | 0.035 |
| *White matter microstructure (n=2867)* | | | | | | | | | | |
| Global FA | 0.013 (-0.001,0.026) | *0.073* | 0.504 | -0.034 (-0.074,-0.006) | **0.021** | 0.168 | 0.000 (-0.032,0.032) | 0.984 | 0.998 | 0.017 |
| Global MD | 0.005 (-0.013,0.021) | 0.616 | 0.788 | 0.025 (-0.003,0.065) | *0.076* | 0.304 | -0.010 (-0.037, 0.022) | 0.604 | 0.995 | 0.025 |
| **Strenuous sports** | | | | | | | | | | |
| *Brain volumes (n=3027)* | | | | | | | | | | |
| Total brain volume | 0.005 (-0.003,0.012) | 0.257 | 0.514 | 0.015 (-0.021,0.052) | 0.409 | 0.630 | 0.013 (-0.017,0.040) | 0.444 | 0.996 | 0.030 |
| Gray matter volume | 0.000 (-0.008,0.008) | 0.944 | 0.944 | 0.010 (-0.026,0.048) | 0.551 | 0.630 | -0.005 (-0.029,0.021) | 0.756 | 0.996 | 0.031 |
| White matter volume | 0.009 (-0.002,0.020) | 0.110 | 0.293 | 0.011 (-0.021,0.044) | 0.480 | 0.630 | 0.026 (-0.008,0.057) | 0.140 | 0.997 | 0.024 |
| White matter hyperintensity | 0.003 (-0.011,0.017) | 0.691 | 0.921 | -0.013 (-0.044,0.015) | 0.337 | 0.630 | -0.014 (-0.029,0.006) | 0.188 | 0.995 | 0.024 |
| Hippocampus volume | 0.011 (0.001,0.019) | **0.026** | 0.104 | 0.036 (0.004,0.070) | **0.027** | 0.216 | 0.011 (-0.019,0.040) | 0.496 | 0.998 | 0.020 |
| Frontal volume | 0.011 (0.002,0.018) | **0.013** | 0.104 | 0.023 (-0.009,0.058) | 0.156 | 0.624 | 0.005 (-0.029,0.039) | 0.770 | 0.995 | 0.036 |

| | | | | | | | | | | |
|---|---|---|---|---|---|---|---|---|---|---|
| *White matter microstructure (n=2867)* | | | | | | | | | | |
| Global FA | 0.001 (-0.012,0.014) | 0.848 | 0.944 | 0.004 (-0.035,0.045) | 0.801 | 0.801 | -0.006 (-0.035,0.024) | 0.717 | 0.968 | 0.020 |
| Global MD | 0.005 (-0.011,0.021) | 0.551 | 0.882 | 0.018 (-0.018,0.064) | 0.271 | 0.630 | 0.005 (-0.024,0.031) | 0.796 | 0.965 | 0.027 |
| **DIY activities** | | | | | | | | | | |
| *Brain volumes (n=3027)* | | | | | | | | | | |
| Total brain volume | 0.008 (-0.003,0.018) | 0.148 | 0.592 | -0.009 (-0.048,0.029) | 0.634 | 0.688 | 0.025 (-0.012,0.053) | 0.217 | 0.997 | 0.026 |
| Gray matter volume | 0.012 (-0.000,0.023) | *0.052* | 0.416 | -0.024 (-0.063,0.014) | 0.214 | 0.342 | 0.017 (-0.018,0.043) | 0.426 | 0.997 | 0.025 |
| White matter volume | 0.002 (-0.015,0.019) | 0.832 | 0.832 | 0.007 (-0.028,0.043) | 0.688 | 0.688 | 0.024 (-0.014,0.058) | 0.224 | 0.998 | 0.018 |
| White matter hyperintensity | -0.003 (-0.022,0.017) | 0.791 | 0.832 | -0.030 (-0.066,-0.000) | **0.047** | 0.094 | 0.037 (-0.001,0.044) | 0.066 | 0.906 | 0.082 |
| Hippocampus volume | -0.005 (-0.015,0.005) | 0.329 | 0.832 | 0.075 (0.037,0.107) | **<0.001** | **<0.001** | 0.018 (-0.015,0.048) | 0.307 | 1.000 | 0.006 |
| Frontal volume | 0.002 (-0.008,0.013) | 0.641 | 0.832 | 0.043 (0.009,0.077) | **0.014** | **0.037** | 0.024 (-0.010,0.055) | 0.181 | 0.995 | 0.007 |
| *White matter microstructure (n=2867)* | | | | | | | | | | |
| Global FA | 0.001 (-0.010,0.013) | 0.821 | 0.832 | 0.042 (0.013,0.081) | **0.007** | **0.028** | -0.020 (-0.049,0.015) | 0.307 | 0.999 | 0.012 |
| Global MD | 0.004 (-0.011,0.018) | 0.619 | 0.832 | -0.013 (-0.056,0.025) | 0.454 | 0.605 | 0.004 (-0.031,0.037) | 0.856 | 0.994 | 0.024 |

breviations: DTI = Diffusion Tensor Imaging, DIY= do-it-yourself, FA=Fractional anisotropy, MD=Mean diffusivity. $\beta_{CL\text{-}1}$ = the cross-lagged path 1, where PA scores at time 1 dict MRI findings at time 2; $\beta_{CL\text{-}2}$ = the cross-lagged path 2, where MRI findings at time 1 predict PA scores at time 2. $\beta_{CS\text{-}Baseline}$ = the cross-sectional association between PA and I within time 1. $p_{FDR}$ = Significant levels, CFI = comparative fit index, RMSEA = root mean square error of approximation. Statistically significant values are shown in bold ($p_{FDR}$ 05). Cross-lagged models were adjusted for age, sex, national origin, educational level, and body mass index.

**Table S4.** Autoregressive associations and complete overview of fit indices of cross-lagged panel models estimating the bidirectional associations between total physical activity and brain structure (brain volumes and global DTI-metrics).

| | Autoregressive | | | | Fit Indices | | | |
|---|---|---|---|---|---|---|---|---|
| | $\beta_{AR\text{-}Physical\ activity}$ | *p* | $\beta_{AR\text{-}Brain}$ | *p* | CFI | TLI | RMSEA | SRMR |
| **Total physical activity** | | | | | | | | |
| *Brain volumes(n=3027)* | | | | | | | | |
| Total brain volume | 0.538 (0.490,0.586) | <0.001 | 0.911 (0.917,0.953) | <0.001 | 0.995 | 0.988 | 0.024 | 0.007 |
| Gray matter volume | 0.554 (0.490,0.587) | <0.001 | 0.906 (0.921,0.964) | <0.001 | 0.995 | 0.989 | 0.023 | 0.007 |
| White matter volume | 0.553 (0.490,0.586) | <0.001 | 0.867 (0.863,0.903) | <0.001 | 0.993 | 0.985 | 0.022 | 0.007 |
| White matter hyperintensity | 0.551 (0.488,0.584) | <0.001 | 0.775 (0.719,1.020) | <0.001 | 0.990 | 0.977 | 0.024 | 0.008 |
| Hippocampus volume | 0.552 (0.489,0.585) | <0.001 | 0.926 (0.908,0.935) | <0.001 | 0.996 | 0.992 | 0.019 | 0.007 |
| Frontal volume | 0.553 (0.489,0.586) | <0.001 | 0.955 (0.964,0.990) | <0.001 | 0.994 | 0.986 | 0.025 | 0.007 |
| *White matter microstructure (n=2867)* | | | | | | | | |
| Global FA | 0.555 (0.488,0.538) | <0.001 | 0.813 (0.924,0.969) | <0.001 | 0.994 | 0.987 | 0.017 | 0.007 |
| Global MD | 0.556 (0.489,0.589) | <0.001 | 0.726 (0.851,0.908) | <0.001 | 0.989 | 0.975 | 0.023 | 0.008 |

Abbreviations: PA= physical activity, FA=Fractional anisotropy, MD=Mean diffusivity. $\beta_{AR\text{-}Physical\ activity}$= the autoregressive coefficient for the PA score, $\beta_{AR\text{-}MRI}$ = the autoregressive coefficient for the MRI score, CFI = comparative fit index, TLI = Tucker-Lewis Index, RMSEA= root mean square error of approximation, SRMR = standardized root mean square residual. Statistically significant values are shown in bold ($p$ <0.05). Cross-lagged models were adjusted for age, sex, national origin, educational level, body mass index, other behaviors (i.e., diet quality and smoking), and other diseases (i.e., hypertension, cancer cardiovascular diseases, diabetes and depression).

**Table S5.** Autoregressive associations and fit indices of cross-lagged panel models estimating the bidirectional associations between physical activity domains and brain structure (brain volumes and global DTI).

| | Autoregressive | | | | Fit Indices | | | |
|---|---|---|---|---|---|---|---|---|
| | $\beta_{AR\text{-Physical activity}}$ | *p* | $\beta_{AR\text{-MRI}}$ | *p* | CFI | TLI | RMSEA | SRMR |
| **Walking for pleasure** | | | | | | | | |
| *Total brain volume (n=3027)* | 0.576 (0.493,0.595) | <0.001 | 0.912 (0.917,0.953) | <0.001 | 0.996 | 0.990 | 0.022 | 0.006 |
| Gray matter volume | 0.576 (0.493,0.596) | <0.001 | 0.906 (0.921,0.963) | <0.001 | 0.996 | 0.991 | 0.021 | 0.006 |
| White matter volume | 0.577 (0.494,0.596) | <0.001 | 0.866 (0.863,0.903) | <0.001 | 0.995 | 0.988 | 0.020 | 0.006 |
| White matter hyperintensity | 0.576 (0.493,0.595) | <0.001 | 0.772 (0.717,1.019) | <0.001 | 0.913 | 0.846 | 0.057 | 0.035 |
| Hippocampus volume | 0.577 (0.494,0.596) | <0.001 | 0.927 (0.903,0.930) | <0.001 | 0.939 | 0.892 | 0.063 | 0.045 |
| Frontal volume | 0.577 (0.494,0.596) | <0.001 | 0.956 (0.964,0.990) | <0.001 | 0.949 | 0.910 | 0.057 | 0.037 |
| *White matter microstructure (n=2867)* | | | | | | | | |
| Global FA | 0.583 (0.494,0.599) | <0.001 | 0.813 (0.925,0.969) | <0.001 | 0.996 | 0.991 | 0.014 | 0.007 |
| Global MD | 0.583(0.494,0.600) | <0.001 | 0.726(0.851,0.908) | <0.001 | 0.992 | 0.982 | 0.020 | 0.007 |
| **Other exercises** | | | | | | | | |
| *Total brain volume (n=3027)* | 0.591 (0.529,0.626) | <0.001 | 0.911 (0.917,0.953) | <0.001 | 0.997 | 0.992 | 0.019 | 0.006 |
| Gray matter volume | 0.591 (0.529,0.626) | <0.001 | 0.906 (0.921,0.964) | <0.001 | 0.997 | 0.993 | 0.019 | 0.006 |
| White matter volume | 0.591 (0.529,0.626) | <0.001 | 0.866 (0.862,0.903) | <0.001 | 0.996 | 0.991 | 0.017 | 0.006 |
| White matter hyperintensity | 0.590 (0.527,0.625) | <0.001 | 0.775 (0.720,1.021) | <0.001 | 0.993 | 0.985 | 0.019 | 0.007 |
| Hippocampus volume | 0.591 (0.529,0.626) | <0.001 | 0.926 (0.908,0.935) | <0.001 | 0.998 | 0.995 | 0.014 | 0.006 |
| Frontal volume | 0.591 (0.529,0.626) | <0.001 | 0.955 (0.964,0.990) | <0.001 | 0.995 | 0.990 | 0.021 | 0.006 |
| *White matter microstructure (n=2867)* | | | | | | | | |
| Global FA | 0.603 (0.530,0.629) | <0.001 | 0.813 (0.924,0.969) | <0.001 | 0.998 | 0.995 | 0.011 | 0.006 |
| Global MD | 0.602 (0.529,0.628) | <0.001 | 0.726 (0.851,0.908) | <0.001 | 0.994 | 0.986 | 0.018 | 0.007 |
| **Strenuous sports** | | | | | | | | |
| *Total brain volume (n=3027)* | 0.512 (0.424,0.582) | <0.001 | 0.911 (0.917,0.953) | <0.001 | 0.995 | 0.990 | 0.021 | 0.007 |
| Gray matter volume | 0.512 (0.424,0.582) | <0.001 | 0.906 (0.921,0.964) | <0.001 | 0.996 | 0.990 | 0.022 | 0.007 |
| White matter volume | 0.511 (0.424,0.581) | <0.001 | 0.866 (0.862,0.902) | <0.001 | 0.994 | 0.988 | 0.020 | 0.007 |
| White matter hyperintensity | 0.512 (0.424,0.581) | <0.001 | 0.776 (0.720,1.021) | <0.001 | 0.990 | 0.979 | 0.023 | 0.008 |
| Hippocampus volume | 0.510 (0.422,0.580) | <0.001 | 0.925 (0.908,0.934) | <0.001 | 0.997 | 0.994 | 0.017 | 0.007 |
| Frontal volume | 0.511 (0.423,0.581) | <0.001 | 0.955 (0.964,0.990) | <0.001 | 0.994 | 0.988 | 0.023 | 0.007 |
| *White matter microstructure (n=2867)* | | | | | | | | |
| Global FA | 0.513 (0.423,0.584) | <0.001 | 0.813 (0.925,0.970) | <0.001 | 0.095 | 0.989 | 0.015 | 0.008 |

| | | | | | | | | |
|---|---|---|---|---|---|---|---|---|
| Global MD | 0.513 (0.422,0.584) | <0.001 | 0.726 (0.851,0.908) | <0.001 | 0.090 | 0.979 | 0.021 | 0.008 |
| **DIY activities** | | | | | | | | |
| *Total brain volume (n=3027)* | 0.401 (0.311,0.453) | <0.001 | 0.911 (0.917,0.953) | <0.001 | 0.997 | 0.993 | 0.017 | 0.006 |
| Gray matter volume | 0.400 (0.311,0.452) | <0.001 | 0.906 (0.921,0.964) | <0.001 | 0.997 | 0.994 | 0.016 | 0.006 |
| White matter volume | 0.401 (0.311,0.452) | <0.001 | 0.866 (0.862,0.903) | <0.001 | 0.997 | 0.992 | 0.015 | 0.006 |
| White matter hyperintensity | 0.400 (0.311,0.452) | <0.001 | 0.773 (0.719,1.020) | <0.001 | 0.903 | 0.828 | 0.056 | 0.035 |
| Hippocampus volume | 0.397 (0.308,0.449) | <0.001 | 0.926 (0.909,0.935) | <0.001 | 0.999 | 0.999 | 0.007 | 0.005 |
| Frontal volume | 0.399 (0.309,0.450) | <0.001 | 0.955 (0.964,0.991) | <0.001 | 0.996 | 0.992 | 0.019 | 0.005 |
| *White matter microstructure (n=2867)* | | | | | | | | |
| Global FA | 0.401 (0.306,0.451) | <0.001 | 0.813 (0.925,0.970) | <0.001 | 0.999 | 0.998 | 0.007 | 0.006 |
| Global MD | 0.401 (0.306,0.452) | <0.001 | 0.726 (0.851,0.908) | <0.001 | 0.993 | 0.984 | 0.017 | 0.007 |

Abbreviations: PA= physical activity, FA=Fractional anisotropy, MD=Mean diffusivity. $\beta_{AR\text{-}Physical\ activity}$= the autoregressive coefficient for the PA score, $\beta_{AR\text{-}MRI}$ = the autoregressive coefficient for the MRI score, CFI = comparative fit index, DIY: do-it-yourself, TLI = Tucker-Lewis Index, RMSEA= root mean square error of approximation, SRMR = standardized root mean square residual. Statistically significant values are shown in bold ($p<0.05$). Cross-lagged models were adjusted for age, sex, national origin, educational level, body mass index, other behaviors (i.e., diet quality and smoking), and other diseases (i.e., hypertension, cancer cardiovascular diseases, diabetes and depression).

**Table S6.** Linear mixed model results for associations that were significant in cross-lagged panel model results (model 2).

| | Brain → Physical activity Time:brain variable | | Cross-sectional Brain variable | | Time | |
|---|---|---|---|---|---|---|
| | β | *Std.error* | $β_{CS}$ | *Std.error* | β | *Std.error* |
| **White matter hyperintensity** | | | | | | |
| Total physical activity | -0.019 | 0.008 | -0.043** | 0.018 | -0.008 | 0.008 |
| DIY activities | -0.021** | 0.010 | -0.010 | 0.019 | -0.026*** | 0.009 |
| **Total brain volume** | | | | | | |
| Walking | 0.017** | 0.007 | 0.003 | 0.020 | 0.023*** | 0.008 |
| **Hippocampal volume** | | | | | | |
| Total physical activity | 0.020** | 0-008 | -0.001 | 0.019 | -0.009 | 0.008 |
| Sports | -0.006 | 0.008 | 0.019 | 0.019 | -0.005 | 0.008 |
| DIY activities | 0.026 *** | 0.009 | 0.017 | 0.020 | -0.026*** | 0.009 |
| **Frontal volume** | | | | | | |
| Domestic work | 0.013 | 0.009 | 0.022 | 0.018 | -0.026*** | 0.009 |
| **Global FA** | | | | | | |
| DIY activities | 0.025** | 0.010 | -0.023 | 0.019 | -0.027*** | 0.010 |
| Other exercises | -0.019** | 0.008 | 0.002 | 0.019 | -0.011 | 0.008 |
| | **Physical activity → Brain Time:brain variable** | | **Cross-sectional Physical activity variable** | | **Time** | |
| | β | *p* | $β_{CS}$ | *p* | β | *p* |
| **Total physical activity** | | | | | | |
| Gray matter volume | 0.004 | 0.004 | 0.004 | 0.014 | -0.095*** | 0.004 |
| **Walking** | | | | | | |
| Total brain volume | -0.012 *** | 0.004 | 0.010 | 0.015 | -0.117*** | 0.003 |
| White matter volume | -0.017*** | 0.004 | 0.016 | 0.017 | -0.100 *** | 0.004 |
| **Sports** | | | | | | |
| Hippocampal volume | 0.010*** | 0.003 | 0.003 | 0.016 | -0.070*** | 0.003 |
| Frontal volume | 0.004 | 0.003 | 0.006 | 0.017 | -0.060*** | 0.003 |

Abbreviations: DIY= do-it-yourself, PA= physical activity. β= the change in physical activity per year increase in follow-up time per one unit of the referring brain variable. $β_{CS}$= the intercept difference of physical activity per one unit of the referring brain variable. Linear mixed models were adjusted for age, sex, national origin, educational level, and body mass index, other behaviors (i.e., diet quality and smoking), and other diseases (i.e., hypertension, cancer cardiovascular diseases, diabetes and depression). Significant codes: $p<0.001$ '***', $p<0.01$ '**', $p<0.05$ '*'.

**Table S7.** Bidirectional associations between total physical activity and brain structure based on cross-lagged panel models, excluding participants who were not able to practice physical activity.

| | Physical activity → Brain | | | Brain → Physical activity | | | Cross-sectional | | Fit measures | |
|---|---|---|---|---|---|---|---|---|---|---|
| | $\beta_{CL-1}$ | $p$ | $p_{FDR}$ | $\beta_{CL-2}$ | $p$ | $p_{FDR}$ | $\beta_{CS-Baseline}$ | $p$ | CFI | RMSEA |
| **Total physical activity** | | | | | | | | | | |
| *Brain volumes (n=2998)* | | | | | | | | | | |
| Total brain volume | -0.001 (-0.010,0.009) | 0.862 | 0.914 | 0.018 (-0.017,0.054) | 0.307 | 0.409 | 0.012 (-0.021,0.040) | 0.536 | 0.995 | 0.024 |
| Gray matter volume | 0.012 (0.001,0.022) | **0.032** | 0.240 | 0.005 (-0.032, 0.043) | 0.779 | 0.779 | -0.002 (-0.030,0.027) | 0.918 | 0.995 | 0.023 |
| White matter volume | -0.014 (-0.029,0.002) | *0.090* | 0.240 | 0.021 (-0.011,0.053) | 0.206 | 0.330 | 0.021 (-0.016,0.054) | 0.283 | 0.993 | 0.022 |
| White matter hyperintensity | -0.009 (-0.026,0.009) | 0.347 | 0.463 | -0.039 (-0.074,-0.013) | **0.005** | **0.020** | -0.049 (-0.067,-0.012) | 0.004 | 0.990 | 0.023 |
| Hippocampus volume | -0.001 (-0.011,0.010) | 0.914 | 0.914 | 0.049 (0.017,0.079) | **0.002** | **0.016** | 0.006 (-0.026,0.036) | 0.758 | 0.996 | 0.019 |
| Frontal volume | 0.009 (-0.001,0.019) | *0.065* | 0.240 | 0.025 (-0.007,0.056) | 0.122 | 0.149 | 0.018 (-0.016,0.049) | 0.324 | 0.993 | 0.025 |
| *White matter microstructure (n=2838)* | | | | | | | | | | |
| Global FA | 0.009 (-0.005,0.022) | 0.204 | 0.408 | 0.030 (-0.001,0.069) | *0.056* | 0.709 | 0.015 (-0.018,0.044) | 0.418 | 0.994 | 0.018 |
| Global MD | 0.009 (-0.008,0.025) | 0.311 | 0.463 | -0.008 (-0.049,0.029) | 0.621 | 0.244 | -0.022 (-0.047,0.014) | 0.283 | 0.988 | 0.024 |

Abbreviations: DTI = Diffusion Tensor Imaging, FA=Fractional anisotropy, MD=Mean diffusivity. $\beta_{CL-1}$ = the cross-lagged path 1, where PA scores at time 1 predict MRI findings at time 2; $\beta_{CL-2}$ = the cross-lagged path 2, where MRI findings at time 1 predict PA scores at time 2. $\beta_{CS-Baseline}$ = the cross-sectional association between PA and MRI within time 1. $p$ = Significant levels, CFI = comparative fit index, RMSEA = root mean square error of approximation. Statistically significant values are shown in bold ($p_{FDR}$ <0.05). Cross-lagged models were adjusted for age, sex, educational level, national origin, and body mass index, other behaviors (i.e., diet quality and smoking) and other diseases (i.e., hypertension, cancer cardiovascular diseases, diabetes and depression).

**Table S8.** Bidirectional associations between total physical activity and brain structure based on cross-lagged panel models, included age quadratic term.

| | Physical activity → Brain | | | Brain → Physical activity | | | Cross-sectional | | Fit measures | |
|---|---|---|---|---|---|---|---|---|---|---|
| | $\beta_{CL-1}$ | $p$ | $p_{FDR}$ | $\beta_{CL-2}$ | $p$ | $p_{FDR}$ | $\beta_{CS-Baseline}$ | $p$ | CFI | RMSEA |
| **Total physical activity** | | | | | | | | | | |
| *Brain volumes (n=3027)* | | | | | | | | | | |
| Total brain volume | -0.004 (-0.014,0.006) | 0.444 | 0.510 | 0.024 (-0.006,0.054) | 0.113 | 0.151 | 0.013 (-0.023,0.048) | 0.503 | 0.916 | 0.094 |
| Gray matter volume | 0.009 (-0.002,0.020) | 0.103 | 0.214 | 0.015 (-0.015, 0.046) | 0.321 | 0.367 | 0.002 (-0.032,0.036) | 0.922 | 0.908 | 0.098 |
| White matter volume | -0.017 (-0.001,-0.017) | **0.038** | 0.214 | 0.025 (-0.005,0.056) | 0.106 | 0.150 | 0.021 (-0.016,0.056) | 0.281 | 0.948 | 0.060 |
| White matter hyperintensity | -0.003 (-0.022,0.015) | 0.718 | 0.718 | -0.040 (-0.072,-0.016) | **0.002** | **0.008** | -0.041 (-0.064,-0.007) | 0.016 | 0.918 | 0.064 |
| Hippocampus volume | -0.004 (-0.015,0.007) | 0.446 | 0.510 | 0.049 (0.019,0.077) | **0.001** | **0.008** | 0.000 (-0.032,0.033) | 0.983 | 0.947 | 0.072 |
| Frontal volume | 0.008 (-0.002,0.018) | 0.107 | 0.214 | 0.030 (-0.000,0.061) | 0.052 | 0.104 | 0.018 (-0.017,0.051) | 0.325 | 0.983 | 0.041 |
| *White matter microstructure (n=2867)* | | | | | | | | | | |
| Global FA | 0.006 (-0.007,0.019) | 0.360 | 0.510 | 0.031 (0.002,0.069) | **0.039** | 0.104 | 0.014 (-0.020,0.043) | 0.461 | 0.965 | 0.042 |
| Global MD | 0.014 (-0.003,0.030) | 0.097 | 0.214 | -0.010 (-0.048,0.024) | 0.510 | 0.510 | -0.015 (-0.045,0.021) | 0.476 | 0.901 | 0.068 |

Abbreviations: DTI = Diffusion Tensor Imaging, FA=Fractional anisotropy, MD=Mean diffusivity. $\beta_{CL-1}$ = the cross-lagged path 1, where PA scores at time 1 predict MRI findings at time 2; $\beta_{CL-2}$ = the cross-lagged path 2, where MRI findings at time 1 predict PA scores at time 2. $\beta_{CS-Baseline}$ = the cross-sectional association between PA and MRI within time 1. $p$ = Significant levels, CFI = comparative fit index, RMSEA = root mean square error of approximation. Statistically significant values are shown in bold ($p_{FDR}$ <0.05). Cross-lagged models were adjusted for age$^2$, sex, educational level, national origin, and body mass index, other behaviors (i.e., diet quality and smoking) and other diseases (i.e., hypertension, cancer cardiovascular diseases, diabetes and depression).
